# Can NLP Detect Loneliness in Electronic Health Records? A Proof-of-Concept Study

**DOI:** 10.64898/2026.04.08.26350462

**Authors:** Tricia Park, Sheida Habibi, Jane Lowers, Abeed Sarker, Selen Bozkurt

## Abstract

Loneliness is clinically important but under-documented in electronic health records (EHRs), posing challenges for secondary use and computational phenotyping. This study evaluated whether natural language processing (NLP) methods can detect and classify loneliness severity from clinical notes. Patients with a loneliness survey (mild, moderate, severe) were identified, and notes within six months prior to the survey were retrieved. An expert-expanded lexicon was applied, and transformer models (RoBERTa, ClinicalBERT, Longformer) were fine-tuned for loneliness severity classification. Large language model-based summarization of social and psychiatric history was also tested as an alternative input representation. Performance was evaluated using accuracy, weighted-F_1_, and per-class F_1_. All models achieved modest accuracy (0.3 to 0.7), and struggled to identify severe loneliness, reflecting sparse and inconsistent documentation even among surveyed patients. While summarization marginally improved accuracy, gains primarily reflected mild predictions. Manual review of 100 social worker notes from severely lonely patients found explicit mentions of loneliness in only two cases, confirming that relevant documentation is exceedingly rare. These findings demonstrate that model performance is constrained by the sparse and inconsistent documentation of loneliness in EHRs, rather than by deficiencies in the modeling approach itself.

## 1. Introduction

Loneliness, defined as the perception of unmet social needs, is a public health concern that greatly affects health outcomes and contributes to mental health crises, and its consequences are consistently detrimental [1,2]. Despite this, loneliness is infrequently and inconsistently documented in electronic health records (EHRs), even where structured fields exist [3]. Unstructured notes may contain relevant signals, but manual review is infeasible at scale [4]. Advances in natural language processing (NLP), including transformer models and large language models (LLMs), offer a path to identify social wellbeing indicators from clinical narratives [5-6].

Prior studies have explored the identification of loneliness in specific clinical contexts (e.g., prostate cancer narratives, long-term care notes), focusing primarily on information extraction through lexicon-based or manually annotated approaches [7-9]. However, these efforts have not addressed the classification of loneliness severity or its representation across diverse note types. In this proof-of-concept study, we test whether modern NLP can (i) detect the presence of loneliness-related content and (ii) grade patient-level severity (mild, moderate, severe). Our objective is to assess the feasibility of automated severity classification and to characterize documentation patterns that constrain model performance, thereby informing future strategies for capturing and modeling psychosocial constructs such as loneliness.

## 2. Methods

With approval from the Emory University Institutional Review Board, we accessed data from the Emory Healthcare EHR system. The study cohort comprised adult patients who completed a spiritual health survey containing a single item on “feelings of loneliness,” with three response options: mild, moderate, and severe. For each patient, we retrieved all clinical notes within six months prior to the earliest survey completion date (index date). In total, 4,290 patients met the inclusion criteria, contributing 112,743 clinical notes spanning common document types (e.g., history and physical (H&P), progress, social service, clinic). These patients were split into training (70%), validation (15%), and testing (15%) subsets to maintain consistency across experiments for comparability.

The survey response served as the patient-level reference label for loneliness severity. Labels were treated as an ordinal outcome (mild < moderate < severe) for aggregation and evaluation; no post-hoc relabeling or binarization was applied. To complement quantitative evaluation, we manually reviewed a random sample of 100 social work notes from patients labeled as severely lonely to assess the frequency and form of explicit loneliness documentation.

We developed an expert-curated lexicon to identify language indicative of loneliness within clinical narratives and to support downstream classification experiments [10]. The initial lexicon consisted of 57 phrases derived from prior work by Zhu et al. [7]. To expand coverage, we (i) identified semantically similar expressions using a text-embedding transformer trained on institutional clinical notes, (ii) generated novel candidate phrases with an LLM (GPT-4o-mini), and (iii) refined candidates through manual review of provider-documented patient quotations retrieved via regular expression queries. Two domain experts of clinical informatics and behavioral health independently reviewed and annotated the terms for contextual relevance. Disagreements were adjudicated by a senior reviewer, and only validated entries were retained. The final lexicon was applied to all notes, and patients without at least one match were excluded from the rest of the study. The notes of the final included patients were used for content analysis, where we investigated the distributions of note type.

We evaluated three transformer-based models with complementary strengths. RoBERTa is a general-domain BERT variant pretrained on large-scale text with dynamic masking and optimized hyperparameters, serving as the baseline [11]. ClinicalBERT is pre-trained on clinical narratives from the MIMIC-III corpus and captures domain-specific terminology and linguistic structures typical of medical documentation [12]. Longformer employs a sparse attention mechanism that enables efficient processing of long clinical notes, reducing information loss due to sequence truncation [13]. We trained models on three note types most relevant to social context: H&P (baseline), social service, and clinic notes. Classification was performed at note level, and patient-level labels were obtained by aggregating predictions using the most severe assigned class. To address lengthy and contextually diffused text, we used an instruction-tuned large language model (gpt-oss 120B) to generate structured summaries of social and psychiatric history from H&P notes. These summaries were aggregated at the patient level and used to fine-tune a RoBERTa classifier under the same patient splits.

## 3. Results

Among 4,290 patients who had completed a spiritual health survey, 3,760 had at least one note that exact-matched our lexicon, with 2,535 (67%) patients indicating mild loneliness, 980 (26%) moderate, and 245 (7%) severe. As seen in Figure 1, coverage of loneliness-related content varied substantially across note types. Progress notes and clinic notes had the most matched patients at about 80% for both, while nursing notes and patient communications had the lowest at under 10%. Across note types, the distributions of severity were relatively consistent, with mild and moderate labels more common than severe labels, which were rarer and often absent in many note types.

**Figure 1.**
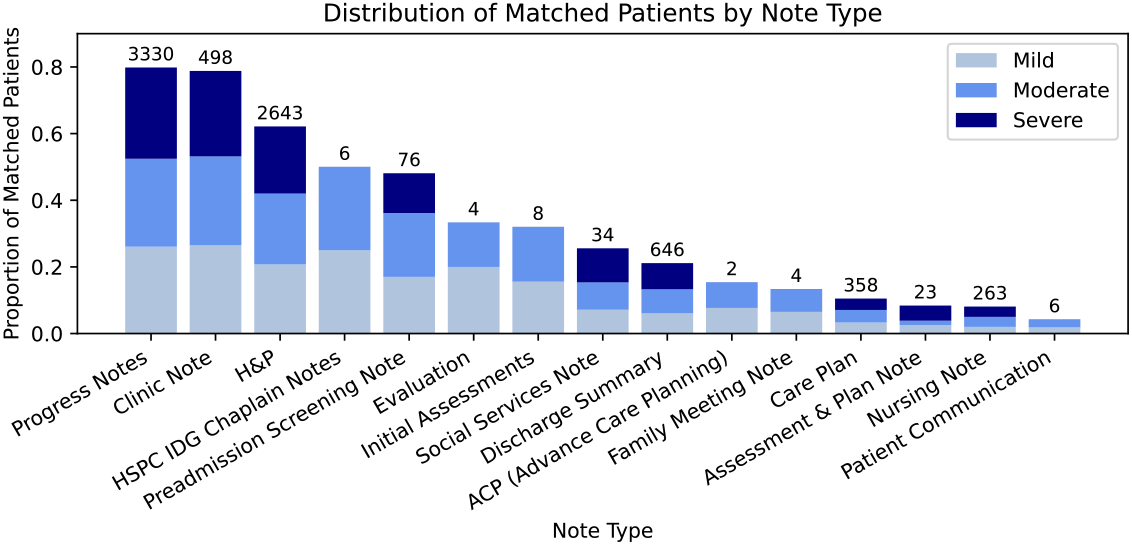
Bar chart showing the distribution of patients with lexicon matches across note types and severity levels. Note types with less than 10 patients were removed. Bar heights represent proportion and annotations atop the bars represent absolute number of matched patients for the respective note types.

Table 1 summarizes the performance of the fine-tuned transformer models across note types. Overall, classification performance was modest across all models and note types, indicating the limited availability of explicit linguistic signals for loneliness in clinical text. Among the models trained on H&P notes, ClinicalBERT achieved the highest F_1_ score (0.565) followed closely by Longformer (0.544), with both outperforming the general-domain RoBERTa baseline (0.447). Although the summarized H&P representation improved RoBERTa’s overall accuracy (0.681), this gain was primarily driven by class imbalance, with the model tending to overpredict the majority (mild) category. Class weighting modestly increased detection of the severe class (F_1_ = 0.113) but reduced overall performance. RoBERTa models trained on social service and clinic notes showed lower and less stable performance (F_1_ = 0.416 and 0.253, respectively), suggesting substantial variability and sparse documentation of loneliness-related concepts across these contexts. Across all experiments, models consistently failed to identify the severe class.

**Table 1.** Classification model performances.

| Input | Model | Accuracy | Weighted F<sub>1</sub> | Mild F<sub>1</sub> | Moderate F<sub>1</sub> | Severe F<sub>1</sub> |
| --- | --- | --- | --- | --- | --- | --- |
| H&P | ClinicalBERT | 0.576 | 0.565 | 0.519 | 0.368 | 0.000 |
| H&P | Longformer | 0.545 | 0.544 | 0.706 | 0.331 | 0.000 |
| H&P | RoBERTa | 0.437 | 0.447 | 0.671 | 0.345 | 0.000 |
| H&P summarized | RoBERTa | 0.681 | 0.554 | 0.810 | 0.015 | 0.000 |
| H&P weighted | RoBERTa | 0.462 | 0.497 | 0.594 | 0.334 | 0.113 |
| Social service | RoBERTa | 0.400 | 0.416 | 0.462 | 0.348 | 0.000 |
| Clinic | RoBERTa | 0.313 | 0.253 | 0.189 | 0.415 | 0.222 |

Model errors primarily stemmed from the absence of explicit loneliness terminology and reliance on indirect or contextual cues. Representative excerpts from notes of patients labeled as severely lonely included statements such as: “Does not have friends or good social support,” “He lives alone and has not been taking care of himself,” “He lives alone, drinks because of boredom. No family or support group locally,” and “Reports stress from [diagnosis], lack of support network; tone of voice was sad and quiet.” A manual review of 100 randomly selected social work notes from these patients identified explicit mentions of loneliness in only two cases, confirming that direct documentation is exceedingly rare even among those known to experience severe loneliness. This finding supports the interpretation that limited documentation, rather than modeling capacity, constitutes the primary barrier to accurate severity classification.

## 4. Discussion and Conclusions

This study evaluated the feasibility of using advanced NLP methods to detect loneliness severity from routine EHR documentation. Despite leveraging transformer models and multiple input strategies, overall performance was modest, reflecting data sparsity rather than model limitations [5]. Loneliness was infrequently and inconsistently documented, often conveyed indirectly through contextual cues (e.g., “lives alone”, “limited social support”) instead of explicit statements. As a result, models tended to overpredict lower-severity categories, mirroring the documentation imbalance observed in the source data.

These findings reveal an expected two-fold bottleneck: limited and inconsistent recording of social and emotional factors, and absence of standardized linguistic or structured indicators of loneliness in EHRs [3-5]. Improving detection will require aligned advances in documentation practices, data standards, and analytic methods, including structured social health fields, and linkage to patient-reported outcomes.

From a modeling perspective, three insights emerge. First, class imbalance and data scarcity constrain learning, especially for severe loneliness. Second, long-context models reduce truncation effects, but they still cannot infer information never documented. Third, summarization increases efficiency but may compress nuanced psychosocial cues, reinforcing the need for representations that balance abstraction with semantic fidelity.

This study has several limitations. Analyses were restricted to a single healthcare system, which may limit generalizability to institutions with different documentation practices. The lexicon-based approach used exact phrase matching, potentially missing linguistic variations or misspellings [14]. In addition, loneliness labels were derived from a spiritual health survey administered to a subset of patients, introducing possible selection bias. Future work should extend analyses to multi-institutional datasets, apply LLMs to capture implicit expressions of loneliness, and explore temporal and note-type specific trends to better understand how loneliness is represented across clinical contexts.

## Data Availability

The data that support the findings of this study are derived from electronic health records (EHRs) containing protected patient information. Due to privacy and institutional regulations, these data are not publicly available. Access may be granted to qualified researchers under a formal data use agreement approved by the relevant institutional review board (IRB) and in compliance with applicable privacy laws and regulations.

## Acknowledgements

Research reported in this manuscript was supported by the National Institute of Mental Health (NIMH) of the National Institutes of Health (NIH) under award numbers 1R21MH139049-01. The content is solely the responsibility of the authors and does not necessarily represent the official views of the NIH.

## References

[1] U.S. Surgeon General. Our epidemic of loneliness and isolation: the U.S. Surgeon General’s advisory on the healing effects of social connection and community. Washington (DC): U.S. Department of Health and Human Services; 2023.

[2] Klinenberg E. Social isolation, loneliness, and living alone: identifying the risks for public health. Am J Public Health. 2016 May;106(5):786–7. doi: 10.2105/AJPH.2016.303166.

[3] Craven CK, Highfield L, Basit M, Bernstam EV, Choi BY, Ferrer RL, et al. Toward standardization, harmonization, and integration of social determinants of health data: a Texas Clinical and Translational Science Award institutions collaboration. J Clin Transl Sci. 2024 Jan 9;8(1):e17. doi: 10.1017/cts.2024.2.

[4] Hatef E, Rouhizadeh M, Tia I, Lasser E, Hill-Briggs F, Marsteller J, et al. Assessing the availability of data on social and behavioral determinants in structured and unstructured electronic health records: a retrospective analysis of a multilevel health care system. JMIR Med Inform. 2019 Aug 2;7(3):e13802. doi: 10.2196/13802.

[5] Bompelli A, Wang Y, Wan R, Singh E, Zhou Y, Xu L, et al. Social and behavioral determinants of health in the era of artificial intelligence with electronic health records: a scoping review. Health Data Sci. 2021 Aug 24;2021:9759016. doi: 10.34133/2021/9759016.

[6] Nerella S, Bandyopadhyay S, Zhang J, Contreras M, Siegel S, Bumin A, et al. Transformers and large language models in healthcare: A review. Artif Intell Med. 2024 Aug;154:102900. doi: 10.1016/j.artmed.2024.102900.

[7] Zhu VJ, Lenert LA, Bunnell BE, Obeid JS, Jefferson M, Halbert CH. Automatically identifying social isolation from clinical narratives for patients with prostate Cancer. BMC Med Inform Decis Mak. 2019 Mar 14;19(1):43. doi: 10.1186/s12911-019-0795-y.

[8] Parmar M, Ma R, Attygalle S, Herath MD, Mueller C, Stubbs B, et al. Associations between recorded loneliness and adverse mental health outcomes among patients receiving mental health-care in South London: a retrospective cohort study. Soc Psychiatry Psychiatr Epidemiol. 2024 Dec;59(12):2155–2164. doi: 10.1007/s00127-024-02663-9.

[9] Rickman S, Fernandez JL, Malley J. Understanding patterns of loneliness in older long-term care users using natural language processing with free text case notes. PLoS One. 2025 Apr 2;20(4):e0319745. doi: 10.1371/journal.pone.0319745.

[10] Bozkurt S, Habibi S, Park T, Blosnich J, Lowers J, Sarker A. ELSDAP resource: detecting loneliness and isolation in EHR narratives [Internet]. 2025 [cited 2025 Oct 16]. Available from: https://osf.io/au8hc

[11] Liu Y, Ott M, Goyal N, Du J, Joshi M, Chen D, et al. RoBERTa: a robustly optimized BERT pretraining approach. 1907.11692 [Preprint]. 2019 [cited 2025 Oct 16]. Available from: https://arxiv.org/abs/1907.11692

[12] Huang K, Altosaar J, Ranganath R. ClinicalBERT: modeling clinical notes and predicting hospital readmission. 1904.05342 [Preprint]. 2019 [cited 2025 Oct 16]. Available from: https://arxiv.org/abs/1904.05342

[13] Beltagy I, Peters ME, Cohan A. Longformer: the long-document transformer. 2004.05150 [Preprint]. 2020 [cited 2025 Oct 16]. Available from: https://arxiv.org/abs/2004.05150

[14] Liu L, Blake V, Barman M, Gallego B, Churches T, Kennedy G, et al. Using natural language processing to extract information from clinical text in electronic medical records for populating clinical registries: a systematic review. J Am Med Inform Assoc. 2026 Feb 1;33(2):484–499. doi: 10.1093/jamia/ocaf176.

